# Measuring Oxygen Access: lessons from health facility assessments in Nigeria

**DOI:** 10.1101/2021.04.21.21255772

**Authors:** Hamish R Graham, Omotayo E Olojede, Ayobami A Bakare, Agnese Iuliano, Oyaniyi Olatunde, Adamu Isah, Adams Osebi, Tahlil Ahmed, Rochelle Ann Burgess, Eric D McCollum, Tim Colbourn, Carina King, Obioma C Uchendu, Adegoke G Falade on behalf of the INSPIRING Project Consortium

**Affiliations:** Centre for International Child Health, Murdoch Children’s Research Institute, University of Melbourne, Royal Children’s Hospital, Parkville, Victoria, Australia; Department of Paediatrics, University College Hospital, Ibadan, Nigeria; Department of Community Medicine, University College Hospital, Ibadan, Nigeria; Department of Global Public Health, Karolinska Institutet, Stockholm, Sweden; Institute for Global Health, University College London, London, UK; Save the Children International, Abuja, Nigeria; Save the Children UK, London, UK; Global Program in Respiratory Sciences, Eudowood Division of Pediatric Respiratory Sciences, Department of Pediatrics, School of Medicine, Johns Hopkins University, Baltimore, USA; Department of Community Medicine, University of Ibadan, Ibadan, Nigeria; Department of Paediatrics, University of Ibadan, Ibadan, Nigeria

**Keywords:** oxygen, pulse oximetry, pneumonia, healthcare providers, health services, management

## Abstract

The COVID-19 pandemic has highlighted global oxygen system deficiencies and revealed gaps in how we understand and measure “oxygen access”. We present a case study on oxygen access from 58 health facilities in Lagos state, Nigeria. We found large differences in oxygen access between facilities (primary vs secondary, government vs private) and describe four key domains to consider when measuring oxygen access.

**Use:** 8/58 (14%) of facilities had a functional pulse oximeter for detecting hypoxaemia (low blood oxygen level) and guiding oxygen care. Oximeters were typically located in outpatient clinics (12/27, 44%), paediatric ward (6/27, 22%), or operating theatre (4/27, 15%), not suitable for children, and infrequently used.

**Availability:** 34/58 (59%) facilities had a functional source of oxygen available on the day of inspection, of which 31 (91%) facilities had it available in a single ward area, typically the operating theatre or maternity ward.

**Cost:** Oxygen was free to patients at primary health centres, when available, but expensive in hospitals and private facilities, with the median cost for 2 days oxygen 13000 ($36 USD) and 27500 ($77 USD) naira, respectively.

**Patient access:** No facilities were adequately equipped to meet minimum oxygen demands for patients. We were unable to determine the proportion of hypoxaemic patients who received oxygen therapy with available data.

We highlight the importance of a multi-faceted approach to measuring oxygen access that assesses access at the point-of-care, and ideally at the patient-level. We propose standard metrics to report oxygen access and describe how these can be integrated into routine health information systems and existing health facility assessment tools.

**SUMMARY BOX:**

- Oxygen access is poorly understood and the most commonly used metrics (e.g. presence of an oxygen source) do not correlate well with actual access to patients.
- Pulse oximetry use is a critical indicator for the quality of oxygen services and may be a reasonable reflection of oxygen coverage to patients with hypoxaemia.
- Oxygen, and pulse oximeter, *availability* must be assessed at the point-of-care in all major service delivery areas, as intra-facility oxygen distribution is highly inequitable.
- Minimum functional requirements for oxygen sources must be assessed, as many oxygen concentrators and cylinders may be present without being in working order.

## BACKGROUND

Oxygen therapy is an essential medicine, required for stabilisation and treatment of severely ill patients with conditions such as COVID-19, pneumonia, and sepsis, and safe anaesthetic care. ^1^ Reliable oxygen services are therefore crucial for every health facility that cares for unwell newborns, children, or adults, and every facility providing obstetric, surgical and post-operative care.

The COVID-19 pandemic has highlighted the importance of hospital oxygen systems and exacerbated existing deficiencies. ^2^ The pandemic has also revealed gaps in how we understand “oxygen access” and the tools we use to measure it. ^3^

In this paper, we use an illustrative case study from Nigeria to describe four domains of data required to meaningfully assess oxygen access. We aim to share lessons learnt about assessing oxygen systems in a variety of facility contexts, to provide guidance for hospital managers, health administrators, policymakers, funders, and supporting non-governmental organizations (NGOs).

### What do we mean by “oxygen access”?

Access to medicines from a health systems perspective involves medication *availability, affordability, quality*, and *rational use*. ^4^ This means that oxygen access requires quality oxygen therapy to be available and affordable to those who need it, when they need it, and used in a safe and rational way.

While these four core domains apply to all essential medicines, oxygen systems are somewhat unique. Medical oxygen must achieve high quality standards – including being ≥85% in purity. For medical oxygen to be safely administered, delivery devices must be appropriate for the particular patient and indication (e.g. nasal cannula, mask with reservoir bag) and guided by the clinical situation and blood oxygen measurements (e.g. SpO_2_, peripheral oxygen saturation from a pulse oximeter). Medical oxygen must be continually available at points-of-care throughout health facilities – from emergency departments, to wards, and operating theatres. Achieving this depends on a range of medical technologies and devices (e.g. oxygen concentrators, cylinders, piping), people (e.g. technicians, nurses/midwives, doctors, administrators) and broader infrastructure and logistic systems (e.g. power supply, maintenance and repair tools and systems) (Figure 1).

**Figure 1.**
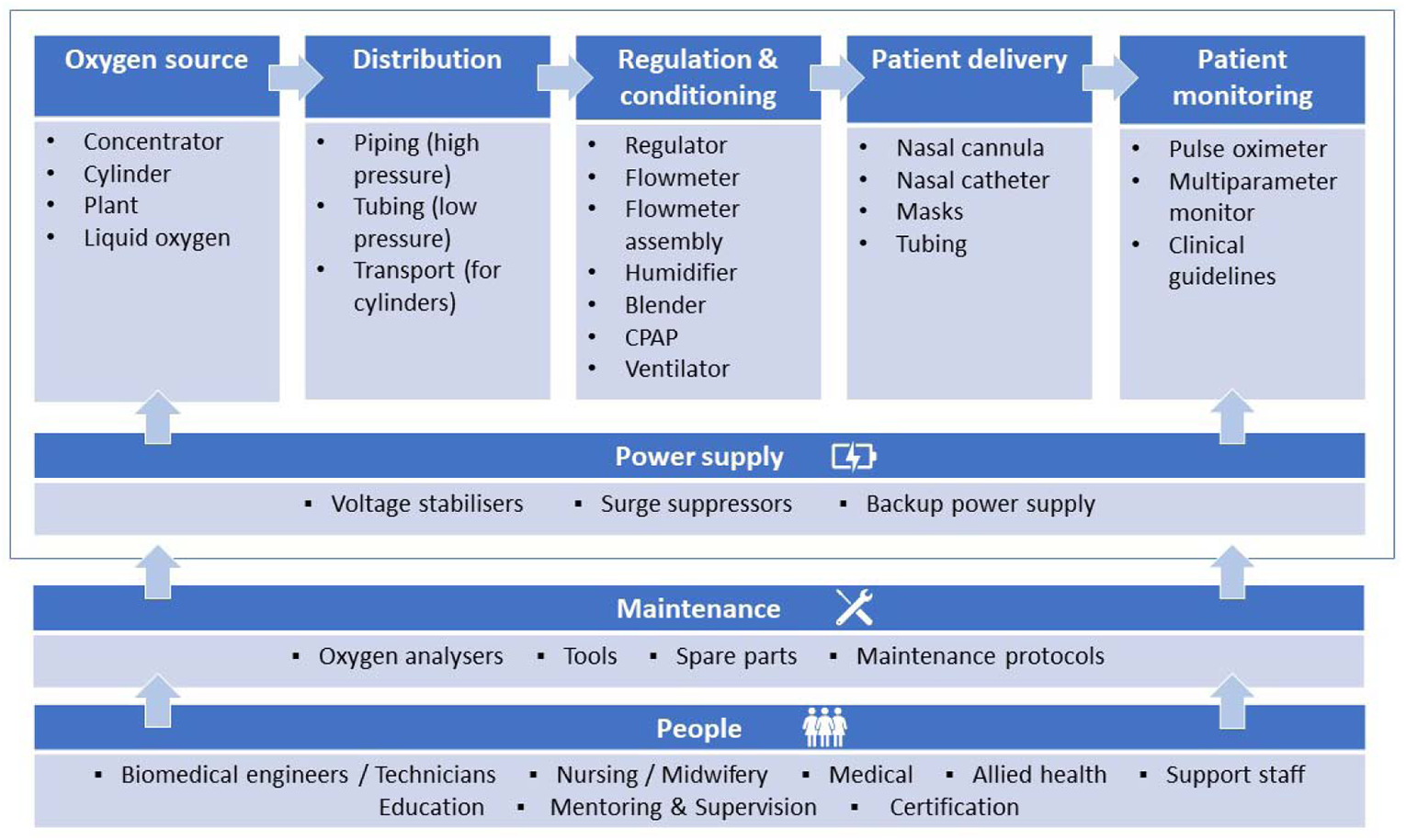
**Hospital oxygen systems require a range of medical devices and other equipment and supplies (Adapted from WHO-UNICEF Technical Specifications and Guidance for Oxygen Therapy Devices)**

Current widely used routine facility survey tools focus on the binary question of “is oxygen equipment available?”. This approach neglects many of these other important dimensions, as well as whether the equipment is functional, what clinical areas it serves, whether healthcare workers can recognise who requires oxygen therapy, or if patients can afford it. As such, this binary approach is a poor reflection of whether patients who need oxygen are actually getting it. ^5^

## Case Study: INSPIRING project, Lagos, Nigeria

Nigeria is a populous lower-middle income country in west Africa with high child (2019: under-five mortality rate 117.2 per 1,000 live births) and maternal mortality ratios (2017: 770 per 100,000 live births)^6^. Lagos is the most populous state, located in the south-west on the Gulf of Guinea. The Lagos population live in urban/peri-urban environments and have lower child mortality rates (50 per 1,000 live births) and poverty levels (1.1% live in severe poverty) compared to Nigeria as a whole. ^7^

Recent studies in Nigeria have shown major deficiencies in existing hospital oxygen systems and highlighted the importance of oxygen in improving pneumonia case management and preventing deaths.^5 8-12^ To better understand the current oxygen capacity and needs of health facilities in Lagos, we surveyed 58 health facilities in Ikorodu local government area (LGA), with an emphasis on paediatric care. We included all government primary health centres (PHCs, n=28), all secondary healthcare facilities (SHFs, n=3) and a random sample of private primary care facilities (n=27/148) (Figure 2). There are no tertiary care facilities in Ikorodu LGA.

**Figure 2.**
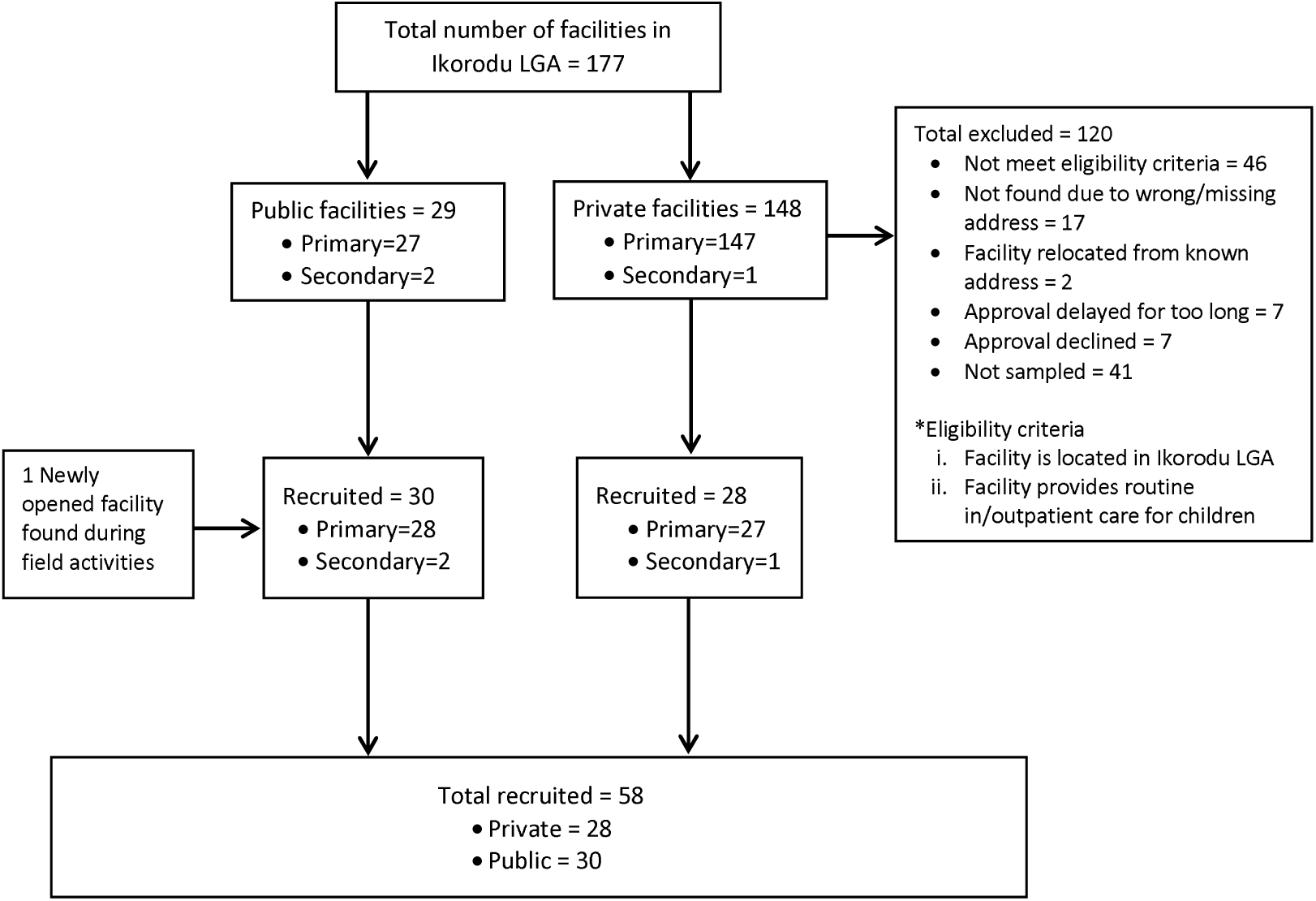
Flow diagram of showing selection of 58 health facilities in Lagos, Nigeria. Notes: Initial identification of facilities conducted in November 2019. We randomly assigned numbers to private facilities then screened and selected until reaching the pre-specified target number for enrolment.

Trained data collectors conducted facility visits between January and August 2020, collecting data from observation, equipment testing and staff surveys using a standardised form, input directly onto tablet computers using CommCare (Dimagi, Cambridge MA, USA). We used Handi+ and UltraMax02 oxygen analysers (Maxtec, Salt Lake City, UT, USA) to test oxygen purity and ProSim SPOT Light (Fluke Corporation, Everett WA, USA) to test pulse oximeter function. We cleaned and analysed data using Stata 15 (StataCorp, College Station TX, USA), and report data descriptively under four broad domains: patient access; oxygen availability; oxygen use; oxygen cost (Figure 3).

**Figure 3.**
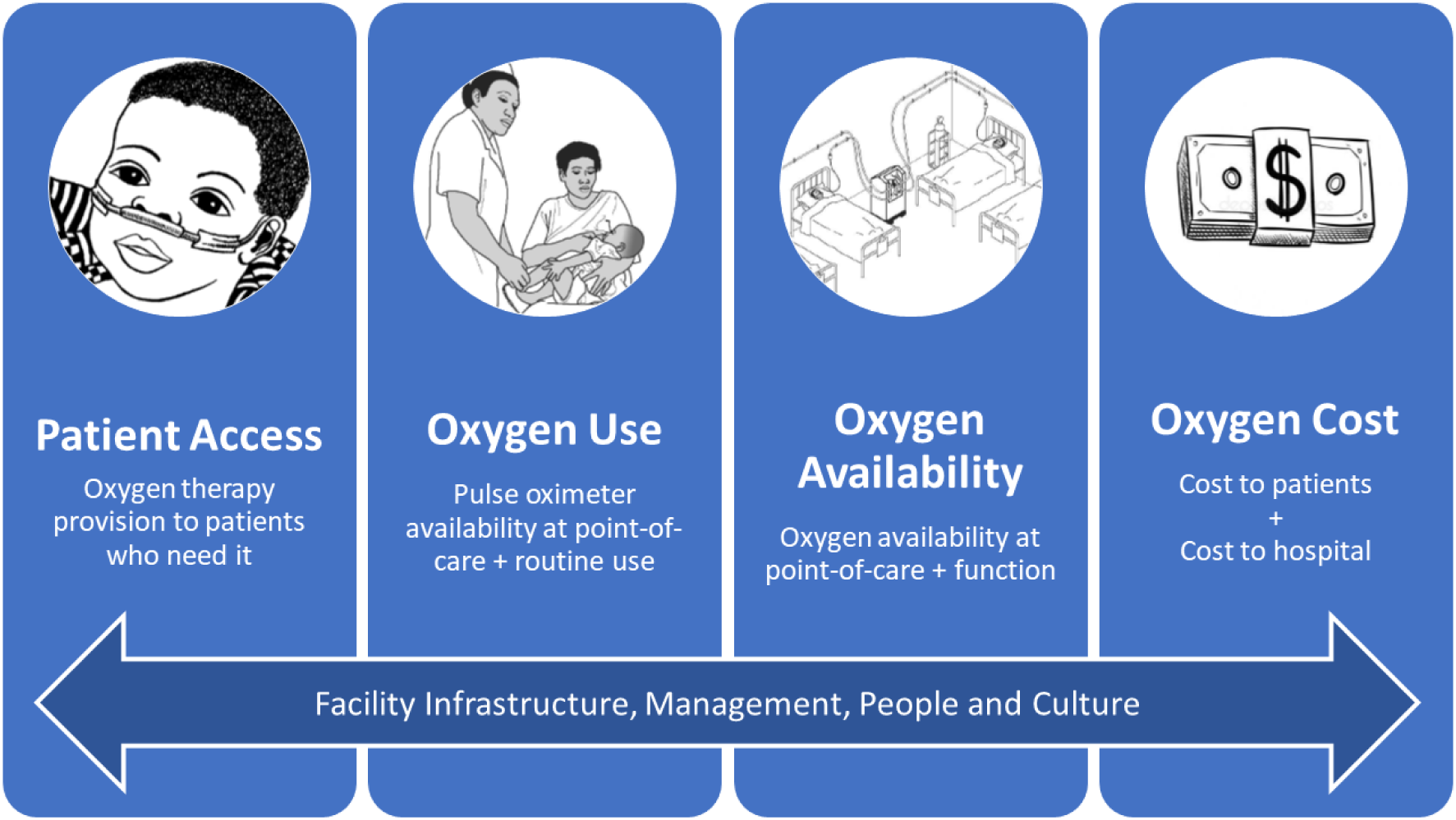
Four domains of data on the quality of “oxygen access”.

### Patient and Public Involvement

This study was conducted to inform the “Integrated Sustainable childhood Pneumonia and Infectious disease Reduction in Nigeria” (INSPIRING) program, implemented by Save the Children to improve child health in Jigawa and Lagos states. INSPIRING involved civil society representatives in co-design activities, including a co-design workshop in April 2019 involving representatives from civil society, local and national government, and professional organisations, together with Save the Children, GSK, and evaluation partners. Selection of the facilities was conducted in partnership with the Ikorodu local government. Community perspectives were sought during the situational analysis but did not contribute substantively to this study.

## Oxygen Use

Pulse oximetry is the key to rational oxygen use, enabling health care workers to accurately detect hypoxaemia (low blood oxygen level), guide oxygen treatment decisions, and subsequently reduce mortality. ^9 13^

### Is a pulse oximeter present and functional?

We located pulse oximeters in all three SHFs–exclusively located in the paediatric wards; none of the government PHCs, and 56% (15/27) of the private primary care facilities. In the private facilities, oximeters were typically located in outpatient clinics (12/27, 44%), paediatric ward (6/27, 22%), or operating theatre (4/27, 15%) (Table 1). The two government hospitals were the only facilities that had paediatric oximetry probes available.

**Table 1.**
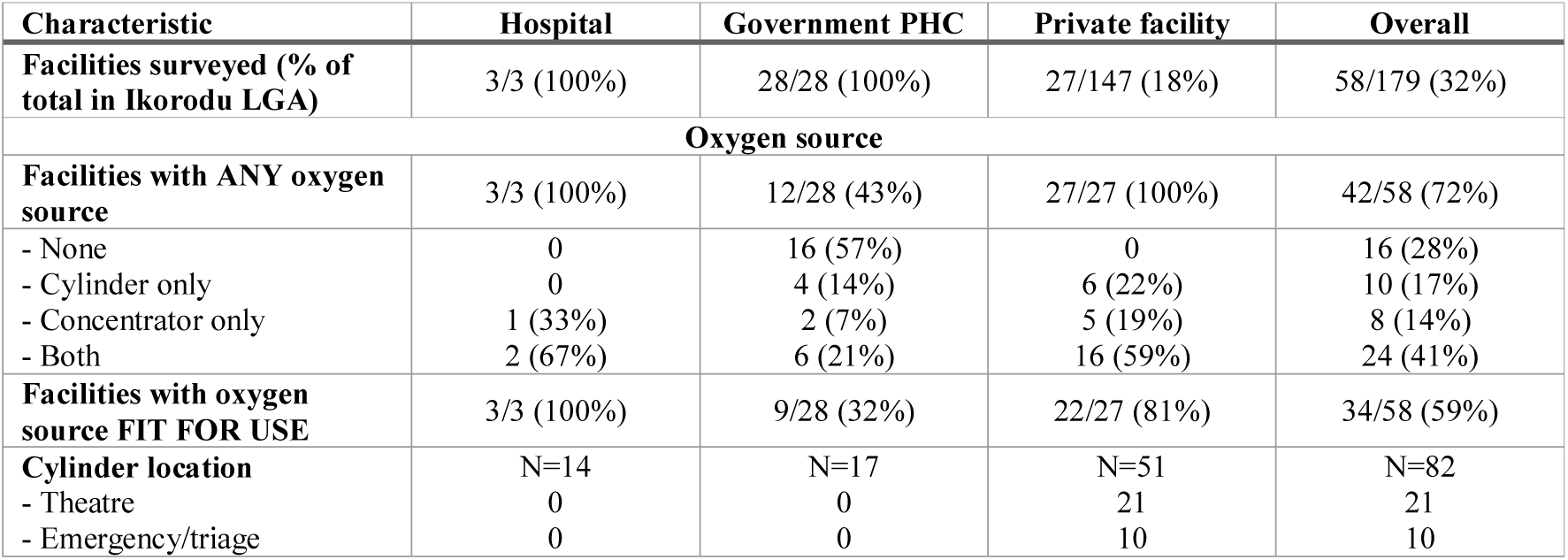
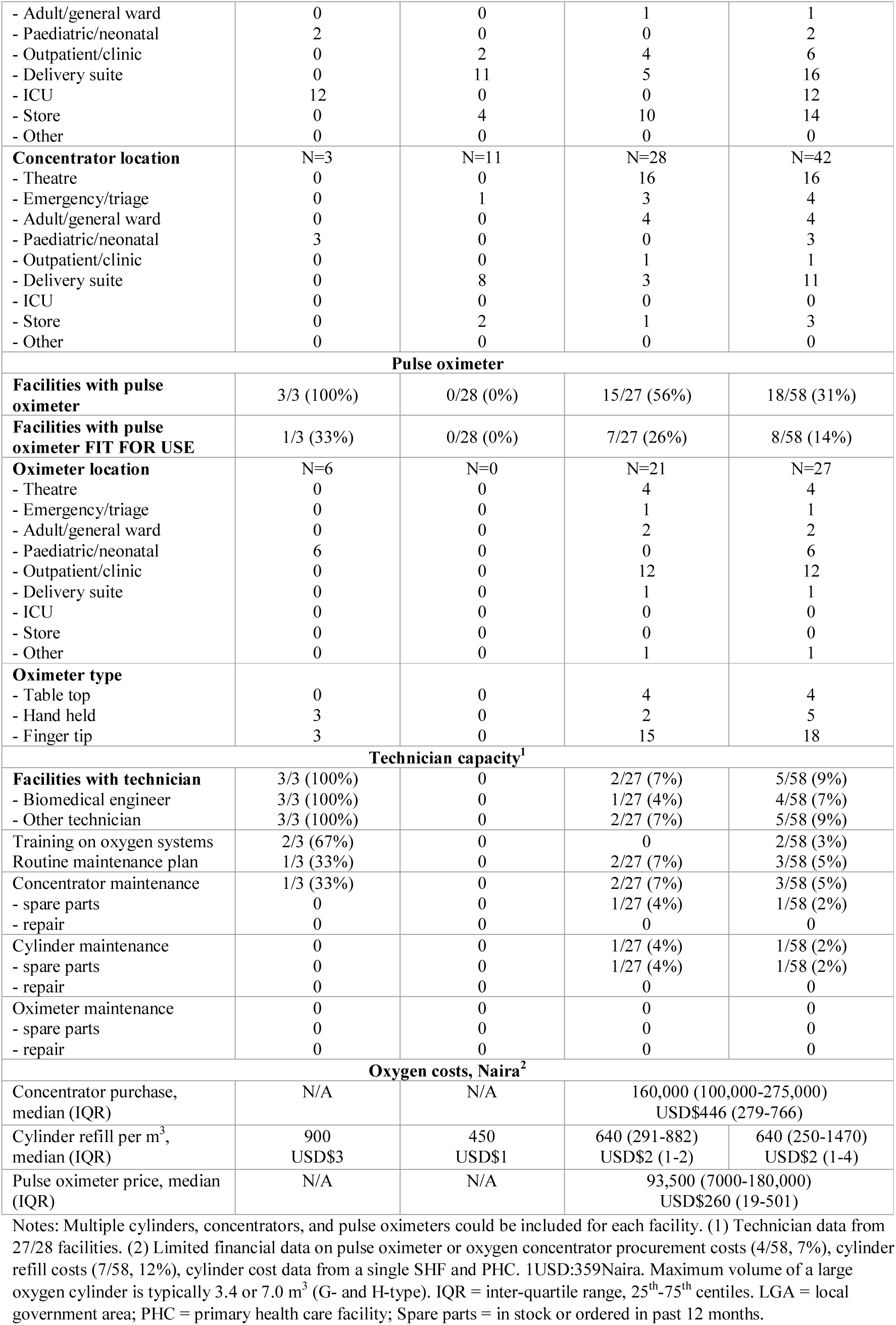
Oxygen access summary data for secondary and primary health facilities in Ikorodu LGA, Lagos, Nigeria.

| Characteristic | Hospital | Government PHC | Private facility | Overall |
| --- | --- | --- | --- | --- |
| **Facilities surveyed (% of total in Ikorodu LGA)** | 3/3 (100%) | 28/28 (100%) | 27/147 (18%) | 58/179 (32%) |
| **Oxygen source** |  |  |  |  |
| **Facilities with ANY oxygen source** | 3/3 (100%) | 12/28 (43%) | 27/27 (100%) | 42/58 (72%) |
| - None | 0 | 16 (57%) | 0 | 16 (28%) |
| - Cylinder only | 0 | 4 (14%) | 6 (22%) | 10 (17%) |
| - Concentrator only | 1 (33%) | 2 (7%) | 5 (19%) | 8 (14%) |
| - Both | 2 (67%) | 6 (21%) | 16 (59%) | 24 (41%) |
| **Facilities with oxygen source FIT FOR USE** | 3/3 (100%) | 9/28 (32%) | 22/27 (81%) | 34/58 (59%) |
| **Cylinder location** | N=14 | N=17 | N=51 | N=82 |
| - Theatre | 0 | 0 | 21 | 21 |
| - Emergency/triage | 0 | 0 | 10 | 10 |
| - Adult/general ward | 0 | 0 | 1 | 1 |
| - Paediatric/neonatal | 2 | 0 | 0 | 2 |
| - Outpatient/clinic | 0 | 2 | 4 | 6 |
| - Delivery suite | 0 | 11 | 5 | 16 |
| - ICU | 12 | 0 | 0 | 12 |
| - Store | 0 | 4 | 10 | 14 |
| - Other | 0 | 0 | 0 | 0 |
| **Concentrator location** | N=3 | N=11 | N=28 | N=42 |
| - Theatre | 0 | 0 | 16 | 16 |
| - Emergency/triage | 0 | 1 | 3 | 4 |
| - Adult/general ward | 0 | 0 | 4 | 4 |
| - Paediatric/neonatal | 3 | 0 | 0 | 3 |
| - Outpatient/clinic | 0 | 0 | 1 | 1 |
| - Delivery suite | 0 | 8 | 3 | 11 |
| - ICU | 0 | 0 | 0 | 0 |
| - Store | 0 | 2 | 1 | 3 |
| - Other | 0 | 0 | 0 | 0 |
| **Pulse oximeter** |  |  |  |  |
| **Facilities with pulse oximeter** | 3/3 (100%) | 0/28 (0%) | 15/27 (56%) | 18/58 (31%) |
| **Facilities with pulse oximeter FIT FOR USE** | 1/3 (33%) | 0/28 (0%) | 7/27 (26%) | 8/58 (14%) |
| **Oximeter location** | N=6 | N=0 | N=21 | N=27 |
| - Theatre | 0 | 0 | 4 | 4 |
| - Emergency/triage | 0 | 0 | 1 | 1 |
| - Adult/general ward | 0 | 0 | 2 | 2 |
| - Paediatric/neonatal | 6 | 0 | 0 | 6 |
| - Outpatient/clinic | 0 | 0 | 12 | 12 |
| - Delivery suite | 0 | 0 | 1 | 1 |
| - ICU | 0 | 0 | 0 | 0 |
| - Store | 0 | 0 | 0 | 0 |
| - Other | 0 | 0 | 1 | 1 |
| **Oximeter type** |  |  |  |  |
| - Table top | 0 | 0 | 4 | 4 |
| - Hand held | 3 | 0 | 2 | 5 |
| - Finger tip | 3 | 0 | 15 | 18 |
| **Technician capacity<sup>1</sup>** |  |  |  |  |
| **Facilities with technician** | 3/3 (100%) | 0 | 2/27 (7%) | 5/58 (9%) |
| - Biomedical engineer | 3/3 (100%) | 0 | 1/27 (4%) | 4/58 (7%) |
| - Other technician | 3/3 (100%) | 0 | 2/27 (7%) | 5/58 (9%) |
| Training on oxygen systems | 2/3 (67%) | 0 | 0 | 2/58 (3%) |
| Routine maintenance plan | 1/3 (33%) | 0 | 2/27 (7%) | 3/58 (5%) |
| Concentrator maintenance | 1/3 (33%) | 0 | 2/27 (7%) | 3/58 (5%) |
| - spare parts | 0 | 0 | 1/27 (4%) | 1/58 (2%) |
| - repair | 0 | 0 | 0 | 0 |
| Cylinder maintenance | 0 | 0 | 1/27 (4%) | 1/58 (2%) |
| - spare parts | 0 | 0 | 1/27 (4%) | 1/58 (2%) |
| - repair | 0 | 0 | 0 | 0 |
| Oximeter maintenance | 0 | 0 | 0 | 0 |
| - spare parts | 0 | 0 | 0 | 0 |
| - repair | 0 | 0 | 0 | 0 |
| **Oxygen costs, Naira<sup>2</sup>** |  |  |  |  |
| Concentrator purchase, median (IQR) | N/A | N/A | 160,000 (100,000-275,000)<br>USD\$446 (279-766) | |
| Cylinder refill per m<sup>3</sup>, median (IQR) | 900<br>USD\$3 | 450<br>USD\$1 | 640 (291-882)<br>USD\$2 (1-2) | 640 (250-1470)<br>USD\$2 (1-4) |
| Pulse oximeter price, median (IQR) | N/A | N/A | 93,500 (7000-180,000)<br>USD\$260 (19-501) | |

Notes: Multiple cylinders, concentrators, and pulse oximeters could be included for each facility. (1) Technician data from 27/28 facilities. (2) Limited financial data on pulse oximeter or oxygen concentrator procurement costs (4/58, 7%), cylinder refill costs (7/58, 12%), cylinder cost data from a single SHF and PHC. 1USD:359Naira. Maximum volume of a large oxygen cylinder is typically 3.4 or 7.0 m<sup>3</sup> (G- and H-type). IQR = inter-quartile range, 25<sup>th</sup>-75<sup>th</sup> centiles. LGA = local government area; PHC = primary health care facility; Spare parts = in stock or ordered in past 12 months.

**Table 2.** Estimated cost of a 3-day admission, or stabilisation care at government PHC, for a child under 5 years of age with severe pneumonia.

| Item | Median cost (IQR) in Naira and USD<sup>1</sup> |  |  |  |
| --- | --- | --- | --- | --- |
|  | Hospital | Government PHC | Private facility | Overall |
| Admission fee | 1500 (0-5000)<br>\$4 (0-14) | N/A | 10,000 (5000-10,000)<br>\$28 (14-28) | 0 (0-7250)<br>\$0 (0-20) |
| Daily bed fee | 0 (0-1500)<br>\$0 (0-4) | N/A | 2000 (1000-2000)<br>\$6 (3-6) | 0 (0-2000)<br>\$0 (0-6) |
| Antibiotics | no data | 300 (280-300)<br>\$1 (1-1) | 1000 (600-1500)<br>\$3 (2-4) | 400 (300-1000)<br>\$1 (1-3) |
| IV and fluids | no data | 200 (200-200)<br>\$1 (1-1) | 1000 (600-1000)<br>\$3 (2-3) | 550 (200-1000)<br>\$2 (1-3) |
| Oxygen | 13,000 (6000-20,000)<br>\$36 (17-56) | 0 (0-0)<br>\$0 (0-0) | 27,500 (3500-96,000)<br>\$77 (10-267) | 3500 (0-36,000)<br>\$10 (0-100) |
| **TOTAL<sup>2</sup>** | **18,500 (7500-29500)**<br>**\$52 (21-82)** | **150 (0-425)**<br>**\$0 (0-1)** | **45,550 (20,250-109,375)**<br>**\$127 (56-305)** | **15,250 (400-53,000)**<br>**\$42 (1-148)** |

Notes: Based on 3 days in hospital, full course of antibiotics and supportive care, 2 days of oxygen therapy. IQR = interquartile range (25<sup>th</sup> and 75<sup>th</sup> centiles); PHC = primary care facility. (1) 1USD:359 Naira (rate 31 Jan 2020); (2) restricted to 34 facilities that had data on oxygen and admission costs. PHC = primary health care facility.

We tested 27 oximeters in 22 ward areas, including 4 desktop, 5 handheld and 18 fingertip oximeters (Table 1, detail in online supplemental appendix). Models varied widely, half (14/27, 52%) had visible CE markings. 22/27 (81%) oximeters turned on, and 19/27 (70%) provided an SpO_2_ reading. Testing with the Fluke device revealed 13/19 (68%) were within +/-2% of the correct SpO_2_ for a simulated healthy person (one read falsely low, five gave no reading). However, only 9/19 (47%) were within +/-3% for a simulated sick person (two read falsely high, 8 gave no reading). Overall, one-third (9/27) of oximeters were demonstrably functional, including 3/4 (75%) desktop, 1/5 (20%) handheld, and 5/18 fingertip (28%) devices – all at private primary care facilities.

### Is pulse oximetry a routine practice?

We found 17/27 (63%) of the oximeters had not been used in the previous day and only five (19%) had been used ≥10 times (online supplemental appendix). Audit of routine case notes for children presenting with acute illness to outpatients in a sub-sample of 12 participating clinics (April-September 2020) showed that pulse oximetry use was low in secondary care facilities (32%, 21/65 patients had SpO2 documented) and negligible in private facilities (2%, 3/177) and PHCs (<1%, 2/608). Thus, despite 31% of facilities having an oximeter, routine use was rare.

### What is the capacity of healthcare workers to provide oxygen therapy?

We conducted knowledge and skills tests for 169 healthcare workers (HCWs) in 56 of the 58 facilities (online supplemental appendix). Participants included doctors, nurses, midwives, health assistants, and community health extension workers with varying levels of experience (median 12 years).

One quarter (40/169, 24%) of HCWs reported training on oxygen, and one-third of these had been trained in the past 5 years (online supplemental appendix). Most HCWs (97/169, 57%) reported experience using oxygen therapy, though only 13 (8%) had administered oxygen in the prior 2-weeks. Experience with oxygen therapy was low in government primary care facilities (2/96, 2%, had used in previous 2-weeks), despite regularly seeing children who required resuscitation (10/96, 10%, had resuscitated a child in the previous 2-weeks).

Knowledge test results showed generally low appreciation of the fundamentals of oxygen therapy, with many respondents lacking the confidence to answer questions – particularly in government primary care facilities (online supplemental appendix). For example, 22% (37/169) of HCWs could identify the core functions of pulse oximetry (i.e. heart rate and SpO_2_), and 28% (48/169) recognised that a child with an SpO_2_ of 87% warranted oxygen therapy.

## Oxygen Availability

### Is an oxygen source present?

We found all SHFs (3/3) and private facilities (27/27), and half of PHCs (12/28, 43%), had an oxygen source in the facility (Table 1). Similar numbers of facilities used oxygen cylinders and oxygen concentrators (34 versus 32), and many facilities had both (24/58, 41%; Table 1).

Most oxygen cylinders (49/82, 60%) and concentrators (27/42, 64%) were in the operating theatre, intensive care unit, or delivery room (online supplemental appendix). Seven facilities had cylinders in store for distribution to wards on demand.

### Is the oxygen source functional?

We visually inspected and tested 42 _*oxygen concentrators*_ in 36 ward areas; 11 could not be fully tested due to power failure (n=10) or active clinical use (n=1). Models varied greatly and one-third (26/42, 62%) had visible CE markings. We found 5/42 (12%) were functional at the time of inspection (Table 1, online supplemental appendix). All used mains power as the primary power source, but only 4/42 (10%) used a voltage stabiliser to protect the device from power fluctuations.

We visually inspected 82 *oxygen cylinders* in 43 ward areas, finding 53/82 (65%) cylinders had a regulator apparatus available (or manifold connection for piped supply) making them fit for use (Table 1, online supplemental appendix). We did not obtain reliable data on leaks, oxygen purity, or pressure.

Overall, 34/58 (59%) facilities had a functional source of oxygen available on the day of inspection, of which 31 (91%) had it available in a single ward area (11 exclusively in the operating theatre, 10 delivery suite, 5 emergency, 2 clinic room, 1 ward, 2 store) (Table 1, online supplemental appendix). We did not evaluate the presence of appropriate oxygen delivery devices.

### What is the technical capacity for oxygen provision?

Five facilities (9%), including all three SHFs, had onsite biomedical engineers or technicians; but only two of these facilities had staff who had been trained on oxygen equipment (Table 1). Three facilities (5%) had routine maintenance schedules for medical equipment and reported performing oxygen concentrator and/or cylinder maintenance in the past 6 months. One facility reported procurement of concentrator and cylinder spare parts in the past year. We did not evaluate technician skills or knowledge.

## Oxygen Cost

### What is the cost of oxygen services to patients?

We obtained patient oxygen cost data from 39/58 (67%) facilities, which variously billed for oxygen using hourly or daily rates, or per admission, cylinder or episode of use. When available, oxygen was typically free to patients at PHCs, but expensive in SHFs and private facilities. The median cost for 2-days oxygen was 13000 Naira (USD $36) and 27500 Naira (USD $77), respectively, accounting for two-thirds of the cost of a 3-day admission for a child with severe pneumonia. This is consistent with previous findings of high out-of-pockets costs for oxygen services in Nigeria^5^ and reflective of Nigeria’s relatively high out-of-pocket health expenditure (2018: 77% of total health expenditure^14^).

### What is the cost of providing oxygen services to facilities?

We found no facilities that included oxygen-related items in their budgets, and few could provide information on pulse oximeter, oxygen concentrator, or cylinder costs. Limited data from 7 facilities suggested median oximeter purchase cost was 93,500 Naira (USD $260), range 7,000-180,000 ($19-500); concentrator purchase cost was 160,000 Naira (USD $446), range 80,000-350,000 ($223-975); and cylinder refill cost was 640 Naira per cubic metre (∼USD $2), range 250-1470 ($1-4) (Table 1).

## Patient Access

### Is oxygen therapy provided to all patients who need it?

Triangulating the data from the 58 health facilities shows clear gaps between need and access with the vast majority of facilities and ward areas lacking a functional oxygen supply and/or pulse oximetry capacity (online supplemental appendix). At best, 10 facilities had oxygen supply adequate to provide minimum services to at least one ward area but only the two government hospital paediatric wards had both oxygen supplies and routine pulse oximetry. Without routine pulse oximetry, the other facilities are likely to miss up to 50% of patients with hypoxaemia even with adequate oxygen supplies. ^8^

The *capacity* to deliver oxygen services does not tell us whether the patients who need oxygen are actually getting it. The clearest metric to measure actual oxygen access to patients is the proportion of patients with hypoxaemia (typically defined as SpO_2_<90%) who are provided with oxygen therapy. We were unable to obtain this data via clinical audit as pulse oximetry coverage was too low and hypoxaemia cannot be accurately determined using clinical signs alone. ^8^ Alternative approaches to measuring oxygen access to patients are using cross-sectional hypoxaemia and oxygen therapy surveys, ^15^ crude counts of patients who receive oxygen, or relying on HCW recall about oxygen supply stock-outs. ^16^ These approaches are problematic and provide a poor indication of whether oxygen is actually reaching those who need it. ^9^

## Discussion

The COVID-19 pandemic has illuminated the need to better measure access to oxygen therapy and stimulated activity to respond. ^3 17^ As part of a comprehensive response to the pandemic, ^18^ the World Health Organization (WHO) has released guidance on oxygen-related biomedical equipment, including how to conduct facility surveys^19^ and technical specifications. ^20-22^

Many other governmental and non-governmental organisations are also working to document oxygen access capacity and needs in health facilities globally – including large scale facility surveys^23 24^ – and provide practical tools to improve access (e.g. UNICEF online repository at https://bit.ly/OxygenResources). ^17^ However, if these efforts count oxygen equipment without assessing actual oxygen availability and patient access, they will overestimate the capacity of current oxygen systems and condemn patients to sub-standard care – particularly those in smaller and more remote facilities.

We tested strategies to obtain more meaningful data on oxygen access, identifying lessons learned that can help HCWs, hospital managers, health administrators, policymakers, and funders who wish to better understand and respond to the oxygen access crisis (Table 3). We included a wide variety of health care facilities in Lagos state, Nigeria and while our specific findings reflect this urban Nigerian environment, the broader lessons about measuring oxygen access are highly generalisable. We focussed on oxygen access metrics that would be most widely applicable and broadly feasible but recognise that some readers will also want additional detail on specific populations (e.g. neonates), services (e.g. CPAP), or supply systems (e.g. cylinder distribution).

**Table 3.**
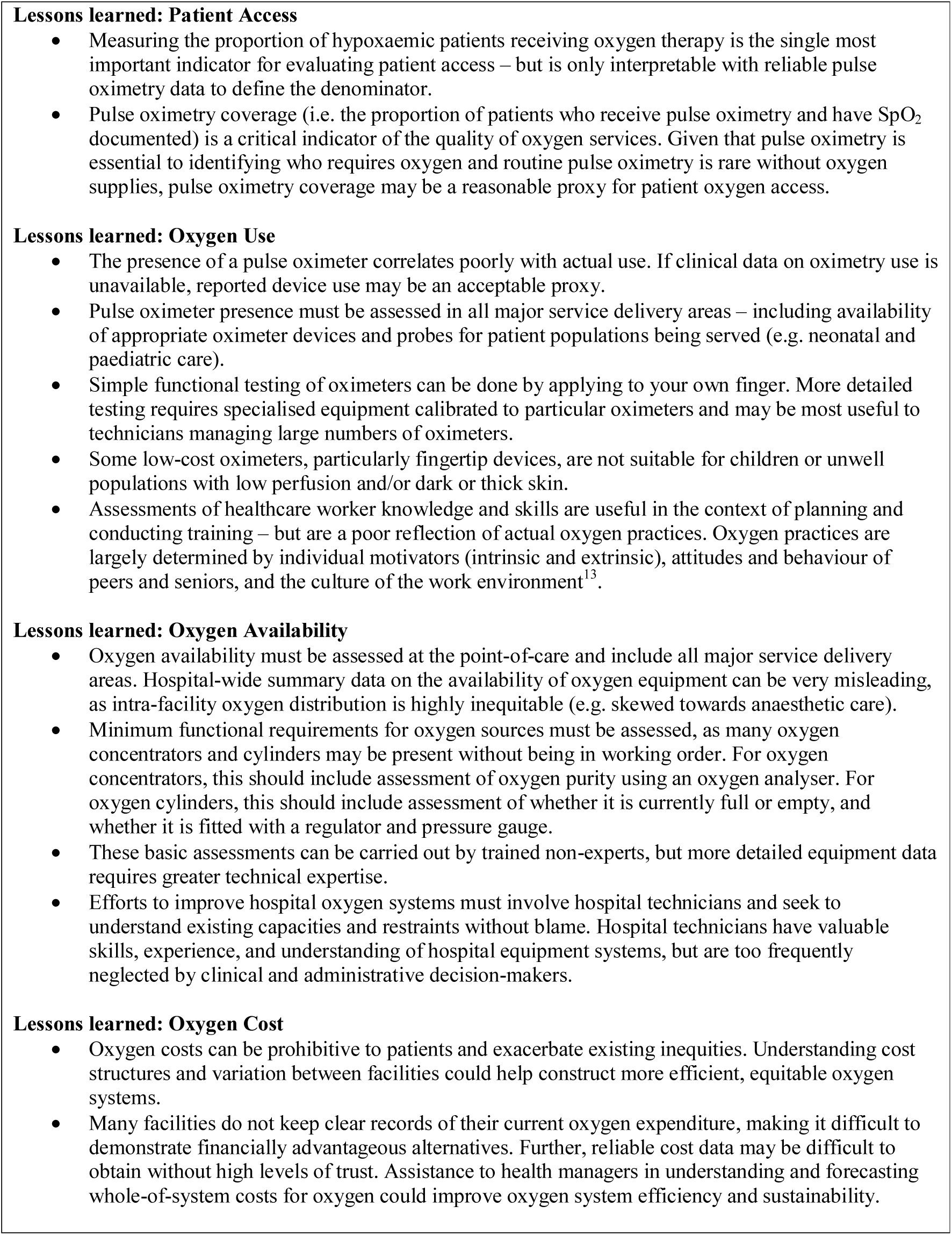
Summary of Lessons Learned on Assessing Oxygen Access.

To aid readers in adapting our approach to local context and needs, we describe four domains of oxygen access that should be considered when seeking to understand and respond to oxygen needs (Figure 3, Table 3, Table 4).

**Table 4.** Summary of most useful indicators of oxygen access.

| **Domain** | **Indicator** | **Potential sources** |
| --- | --- | --- |
| **Patient Oxygen Access** | ***Proportion of patients with hypoxaemia receiving oxygen therapy.***<br>*Additional considerations:*<br>- equity between populations (hospital department, age, sex, ethnic background, socioeconomic status)<br>- rational use (oxygen provision with versus without indication)<br>- flow rates and duration of therapy (useful for quantification of oxygen need and quality improvement) | Clinical audit, or routine health information system |
| **Oxygen Use** | ***Proportion of acutely unwell patients screened using pulse oximetry.***<br>*Additional considerations:*<br>- pulse oximeter presence and function<br>- oxygen clinical guideline<br>- equity between populations (hospital department, age, sex, ethnic background, socioeconomic status)<br>- training and skills of healthcare workers | Clinical audit, routine health information system, physical inspection + device testing, HCW survey |
| **Oxygen Availability** | ***Proportion of wards/departments with functional oxygen source and delivery equipment.***<br>*Additional considerations:*<br>- oxygen source presence and function (including oxygen purity testing for oxygen concentrators)<br>- delivery devices (cannula, mask, CPAP, flowmeter assembly, etc.)<br>- maintenance personnel (local capacity and access to expert help)<br>- equipment inventory and preventive maintenance schedules<br>- spare parts: access and cost | Physical inspection + device testing, technician survey |
| **Oxygen Cost** | ***Daily cost of oxygen therapy to patients***<br>***Annual expenditure on oxygen equipment and maintenance/repairs.***<br>*Additional considerations:*<br>- equity and non-payment action (hospital department, age, poor)<br>- whole of life cycle (capital and operating costs): oxygen source; refill cost; spare parts and maintenance; power; consumables. | Patient accounts and general finance ledger, administrator survey |

At the centre is *patient oxygen access* – evaluating whether patients who need oxygen actually receive it. Evaluating patient oxygen access requires clinical data on *pulse oximetry and oxygen use*. Recent studies from Nigeria and Malawi have shown that without routine pulse oximetry to guide oxygen therapy, only around 20% of hypoxaemic patients receive oxygen – even if adequate oxygen supplies are available. ^9 15^ Given that pulse oximetry is essential to identifying who requires oxygen and routine pulse oximetry is rare without oxygen supplies, ^5 13^ pulse oximetry coverage (i.e. the proportion of patients who receive pulse oximetry and have SpO_2_ documented) may be a reasonable proxy for patient oxygen access.

High level metrics on *pulse oximetry and oxygen use* can be sourced via clinical audit but are ideally integrated into health information systems and are essential for health service planning (Table 4). More detailed data, including who receives oxygen-related care, where, with what, and for how long, would be desirable for hospital-level quality improvement activities and improving oxygen system efficiencies.

While measures of *oxygen availability* are important, existing surveys which contain this metric often fail to assess functionality of equipment or address intra-hospital distribution, missing huge deficiencies in oxygen availability at the point of care. ^5 12^ Basic assessments of equipment functionality and distribution are not difficult and could be integrated into existing surveys such as the Service Provision Assessment (SPA) and Service Availability and Readiness Assessment (SARA) (Table 4). More detailed data on equipment types, locations, and access to spare parts is useful for hospital-level decision-making and efficiency – particularly in facilities providing a wider range of respiratory support services (e.g. CPAP, mechanical ventilation) across multiple service areas.

Measures of *oxygen cost* are important for planning purposes but were challenging to obtain in practice. We propose metrics that address oxygen costs to patients, crucial in contexts where out-of-pocket health expenditure is high, and oxygen costs to facilities/governments obtained from financial records and surveys (Table 4). However, we recognise the difficulties in obtaining sensitive financial information and the limitations of relying on administrator or technician recall in the absence of reliable financial records.

Finally, our case study highlights opportunities for priority improvements in existing oxygen systems. First, pulse oximeters are low-cost devices that were grossly lacking and could substantially improve rational use of existing oxygen supplies. Second, while many devices were not fit for use, support for technicians could likely rehabilitate some of these and prevent future growth of equipment graveyards (e.g. Open O2 in Malawi, www.openo2.org). Third, many devices did not have CE or similar quality control markings, highlighting the ongoing challenge of inappropriate equipment donation and procurement of consumer devices for medical purposes. Fourth, the differences between government and private facilities highlights that oxygen access is fundamentally an issue of equity, and efforts must include smaller, poorer, and more rural facilities that care for the most at-risk populations.

## Conclusion

Data on oxygen access is lacking and makes efforts to improve oxygen systems for COVID-19 and beyond clumsy and inefficient. We propose four data domains to assess oxygen systems: patient access; use; availability; and cost. Measuring these four domains of oxygen access will support local, regional, and global efforts to provide life-saving oxygen to those in need.

## Data Availability

De-identified data is available on request from the authors.

## Authors’ contributions

CK, HG, EDM, AI, AAB, AGF, TC conceived of the study and TC, CK and AGF are grant holders. CK, TC, HG, EDM, AGF, RB, AAB initiated the study design and OEO and OO led data collection and implementation. Data analysis was conducted by HG. The manuscript was drafted by HG, with input from OEO, AAB and CK. All authors contributed to revisions and approved the final manuscript.

## Ethics approval

This study has received ethical approval from the Research Ethics Committee at: University of Ibadan (REF UI/EC/19/0551), Lagos State (REF LS/PHCB/MS/1128/VOL.V1/005), and University College London (REF 3433/005).

