## supplemental material for "Measuring Oxygen Access: lessons from health facility assessments in Nigeria"

**Supplemental table 1**: Healthcare worker knowledge and experience with oxygen, pulse oximetry and related clinical practices

**Supplemental table 2**: Results from testing of pulse oximeter in 58 health facilities in Ikorodu local government area, Lagos state, Nigeria

**Supplemental table 3**: Results from testing of oxygen concentrators in 58 health facilities in Ikorodu local government area, Lagos state, Nigeria

**Supplemental table 4**: Results from testing of oxygen cylinders in 58 health facilities in Ikorodu local government area, Lagos state, Nigeria

**Supplemental table 5**: Triangulation of pulse oximetry and oxygen access to ward areas in 58 health facilities in Ikorodu local government area, Lagos state, Nigeria

### Supplemental table 1: Healthcare worker knowledge and experience with oxygen, pulse oximetry and related clinical practices

| General characteristics | Secondary health facility | Government PHC | Private PHC | Overall |
|---|---|---|---|---|
| Facilities | N=1[1] | N=28 | N=27 | N=56 |
| Participants | 9 | 96 | 64 | 169 |
| Sex, F:M (% female) | 8:1 (89%) | 91:5 (95%) | 50:14 (78%) | 149:20 (88%) |
| Role<br>- doctor<br>- nurse/midwife<br>- other | <br>2 (22%)<br>2 (22%)<br>5 (56%) | <br>5 (5%)<br>49 (51%)<br>42 (44%) | <br>16 (25%)<br>37 (58%)<br>11 (17%) | <br>23 (14%)<br>88 (52%)<br>58 (34%) |
| Median years of work<br>- at any health facility<br>- at this facility | <br>7 (3-15)<br>1 (1-3) | <br>15 (6-29)<br>1 (1-2) | <br>8 (4-15)<br>1 (1-8) | <br>12 (5-25)<br>2 (1-3) |
| Training<br>- IMCI<br>- ICCM<br>- ETAT<br>- Oxygen<br>- Pulse oximetry<br>- CPAP<br>- Resuscitation<br>- Infection control<br>- Baby friendly initiative | <br>2 (22%)<br>0 (0%)<br>1 (11%)<br>1 (11%)<br>0 (0%)<br>1 (11%)<br>4 (44%)<br>4 (44%)<br>5 (56%) | <br>62 (65%)<br>4 (4%)<br>5 (5%)<br>9 (9%)<br>4 (4%)<br>5 (5%)<br>27 (28%)<br>41 (43%)<br>35 (36%) | <br>21 (33%)<br>7 (11%)<br>13 (20%)<br>30 (47%)<br>27 (42%)<br>15 (23%)<br>35 (55%)<br>40 (63%)<br>31 (48%) | <br>85 (50%)<br>11 (7%)<br>19 (11%)<br>40 (24%)<br>31 (18%)<br>21 (12%)<br>66 (39%)<br>85 (50%)<br>71 (42%) |
| **Child pneumonia care** | | | | |
| Ever provided<br>- antibiotic for pneumonia<br>- IV antibiotic<br>- oxygen therapy<br>- resuscitate child | <br>7 (78%)<br>6 (67%)<br>7 (78%)<br>7 (78%) | <br>70 (73%)<br>39 (41%)<br>32 (33%)<br>67 (70%) | <br>56 (88%)<br>57 (89%)<br>58 (91%)<br>59 (92%) | <br>133 (79%)<br>102 (60%)<br>97 (57%)<br>130 (79%) |
| Past 2 weeks provided<br>- antibiotic for pneumonia<br>- IV antibiotic<br>- oxygen therapy<br>- resuscitate child | <br>5 (56%)<br>4 (44%)<br>3 (33%)<br>3 (33%) | <br>35 (36%)<br>8 (8%)<br>2 (2%)<br>10 (10%) | <br>16 (25%)<br>11 (17%)<br>9 (14%)<br>15 (23%) | <br>56 (33%)<br>23 (14%)<br>13 (8%)<br>28 (17%) |
| **Oxygen knowledge** | | | | |
| Mean score (95% CI)<br>- Total score (max. 40)<br>- Yes/No (max. 20)<br>- Scenarios[2] (max. 20) | <br>17.8 (12.1-23.5)<br>10.0 (6.7-13.3)<br>7.8 (4.4-11.2) | <br>6.7 (5.2-8.3)<br>4.4 (3.4-5.3)<br>2.3 (1.6-3.1) | <br>14.9 (13.3-16.4)<br>10.9 (9.9-11.9)<br>4.0 (3.1-4.9) | <br>10.4 (9.1-11.6)<br>7.1 (6.3-8.0)<br>3.3 (2.6-3.9) |
| Sample questions<br>- Correctly identify that pulse oximeters provide heart rate, $SpO_2$ and not blood pressure or respiratory rate[3] | 3 (33%) | 7 (7%) | 27 (42%) | 37 (22%) |
| - Correctly identify that a 2-year-old child with fast breathing and $SpO_2$ of 87% should be started on oxygen | 6 (67%) | 18 (19%) | 24 (38%) | 48 (28%) |
| - Correctly identify that a small newborn baby with $SpO_2$ 99% on oxygen should have the oxygen flowrate reduced | 3 (33%) | 11 (11%) | 18 (28%) | 32 (19%) |

Notes: CI = confidence interval; CPAP – continuous positive airway pressure; ETAT – emergency triage, assessment, and treatment; ICCM – integrated community case management; IMCI = integrated management of childhood illness; IQR = inter-quartile range, 25th to 75th centiles; $SpO_2$ – peripheral oxygen saturation. (1) Two secondary health facilities did not do the knowledge test; (2) 5-option best answer scenario with pulse oximetry result displayed; (3) Composite from 4 individual true/false questions.

**Supplemental table 2: Results from testing of pulse oximeter in 58 health facilities in Ikorodu local government area, Lagos state, Nigeria**

| Facility | Device | Type | Location | Usage (times in prior day) | Oximeter Function | | | |
|---|---|---|---|---|---|---|---|---|
| | | | | | Turns on | Fluke test 1 ($SpO_2$ 95%) | Fluke test 2 ($SpO_2$ 85%) | Fit for use |
| **Secondary health facilities** | | | | | | | | |
| SHF 1 | BCI* | Handheld | Ward (child) | 10 | Y | - | - | N[3] |
| SHF 2 | BCI* | Handheld | Ward (child) | 0 | N | - | - | N |
| | Oxi-Go | Finger tip | Ward (child) | ~100 | Y | 96 | 90 | N |
| | Mindray* | Handheld | Ward (child) | 0 | Y | - | - | N[3] |
| SHF 3 | "Medi Industries" | Finger tip | Ward (child) | 2 | Y | 96 | 86 | Y |
| | "Pediatric" | Finger tip | Ward (child) | 0 | Y | 93 | NR | N |
| **Private primary care facilities** | | | | | | | | |
| PRV 1 | unknown | Finger tip | Clinic | 0 | N | - | - | N |
| | unknown* | Finger tip | Clinic | 0 | N | - | - | N |
| | Contec | Desktop | Theatre | 0 | N[1] | - | - | N[1] |
| PRV 4 | Edan* | Handheld | Clinic | 30 | Y[2] | NR | NR | ? |
| | Edan* | Desktop | Theatre | 5 | Y | 96 | 87 | Y |
| PRV 6 | Drive | Finger tip | Clinic | 50 | Y[2] | NR | NR | ? |
| | Contec* | Finger tip | Emergency | 0 | Y[2] | NR | NR | ? |
| PRV 7 | Edan* | Desktop | Theatre | 0 | Y | 96 | 88 | Y |
| PRV 8 | unknown | Finger tip | Ward (general) | 20 | Y[2] | NR | NR | ? |
| PRV 9 | unknown | Finger tip | Clinic | 0 | Y | 96 | NR | N |
| PRV 10 | Schiller* | Handheld | Other | 0 | Y | 96 | 86 | Y |
| PRV 14 | Ana Wiz* | Finger tip | Clinic | 0 | Y[2] | NR | NR | ? |
| PRV 16 | EcoMed | Finger tip | Clinic | 0 | Y | 96 | 85 | Y |
| PRV 18 | Blue Jay | Finger tip | Clinic | 0 | Y | 95 | 85 | Y |
| PRV 19 | FaceLake | Finger tip | Clinic | 4 | Y | - | - | N[3] |
| | FaceLake | Finger tip | Clinic | 0 | N | - | - | N |
| PRV 22 | Promise Technology* | Finger tip | Clinic | 5 | Y | 96 | NR | N |
| PRV 24 | "Fabrication Enterprises"* | Finger tip | Ward (general) | 3 | Y | 95 | 85 | Y |
| PRV 25 | "Medline" | Finger tip | Delivery | 0 | Y | 97 | 92 | N |
| PRV 27 | Datascope* | Desktop | Theatre | 0 | Y | 96 | 84 | Y |
| | "iHealth"* | Finger tip | Clinic | 0 | Y | 96 | 87 | Y |
| **TOTAL** | | | | | **22/27 (81%)** | **14/27 (52%)** | **9/27 (33%)** | **9/27 (33%)** |

Notes: Fluke test 1 simulate normal person, set to 95% with good perfusion. Fluke test 2 simulate sick person, set to 85% with reduced perfusion. Considered "pass" if within +/-3%. (1) unable to test due to power outage; (2) unable to test with Fluke device; (3) missing probe or other defect preventing use. NR = no result.

**Supplemental table 3: Results from testing of oxygen concentrators in 58 health facilities in Ikorodu local government area, Lagos state, Nigeria**

| Facility | Device type | Location | Concentrator Function[1] | | | | | | |
|---|---|---|---|---|---|---|---|---|---|
| | | | Turns on | Gas flow | Air | 22-49% | 50-84% | ≥85% | Fit for use |
| **Secondary health facilities** | | | | | | | | | |
| SHF 1 | "LuFaith Y007-3"* | Ward (child) | Y | Y | | ○ | | | N |
| SHF 2 | DeVilbiss* | Ward (child) | N | N | | | | | N |
| SHF 3 | unknown | Ward (child) | Y | Y | | ○ | | | N |
| **Private primary care facilities** | | | | | | | | | |
| PRV 1 | Longfe+B2+E6:M41 | Theatre | N[2] | - | | | | | N[2] |
| PRV 2 | Weinmann | Theatre | Y | N | | | | | N |
| PRV 4 | Longfei* | Theatre | Y | Y | ○ | | | | N |
| PRV 4 | Draeger* | Emergency | Y | N | | | | | N |
| PRV 4 | Longfei* | Delivery | Y | Y | ○ | | | | N |
| PRV 6 | unknown "7F-3" | Emergency | Y | Y | | | ○ | | N |
| PRV 7 | unknown | Theatre | Y | Y | ○ | | | | N |
| PRV 8 | "Leap Medical 7F-3" | Ward (general) | Y | Y[4] | | | | | ?[4] |
| PRV 10 | "Elgil LFY-I-3A-W"* | Delivery | Y | Y | ○ | | | | N |
| PRV 10 | "Globe Health 7F-3" | Ward (general) | Y | Y | | | | ○ | Y |
| PRV 12 | Philips* | Delivery | N[2] | - | | | | | N[2] |
| PRV 13 | Invacare | Emergency | Y | Y | | | | ○ | Y |
| PRV 13 | Longfei* | Theatre | N | N | | | | | N |
| PRV 14 | Microfield* | Ward (general) | N[2] | - | | | | | N[2] |
| PRV 14 | Microfield* | Ward (general) | N[3] | N | | | | | N |
| PRV 15 | Zhengzhou Olive* | Theatre | Y | Y | | | | ○ | Y |
| PRV 17 | "Shulte-Deme"* | Store | Y[2] | - | | | | | N[2] |
| PRV 18 | DeVilbiss* | Theatre | Y[2] | - | | | | | N[2] |
| PRV 19 | Microfield* | Theatre | Y[2] | - | | | | | N[2] |
| PRV 20 | "Globe Health"* | Clinic | Y | Y | ○ | | | | N |
| PRV 21 | Puritan Bennett | Theatre | N | N | | | | | N |
| PRV 22 | Philips* | Theatre | N[2] | - | | | | | N[2] |
| PRV 20 | DeVilbiss* | Theatre | N[2] | - | | | | | N[2] |
| PRV 23 | Jiangsu Folee* | Theatre | Y | Y | ○ | | | | N |
| PRV 23 | Jiangsu Folee* | Theatre | Y | Y | ○ | | | | N |
| PRV 24 | unknown* | Theatre | N | N | | | | | N |
| PRV 26 | Airsep* | Theatre | N | N | | | | | N |
| PRV 27 | Invacare | Theatre | Y | Y | ○ | | | | N |
| **Government primary health care facilities** | | | | | | | | | |
| PHC 1 | "Axiom" | Delivery | Y | N | | | | | N |
| PHC 3 | "MA-Donax"* | Delivery | N[2] | - | | | | | N[2] |
| PHC 11 | Microfield* | Delivery | N | N | | | | | N |
| PHC 14 | unknown | Delivery | Y | N | | | | | N |
| PHC 14 | unknown | Delivery | Y | Y | ○ | | | | N |
| PHC 16 | unknown | Store | Y | N | | | | | N |
| PHC 16 | unknown | Store | Y | Y | | | | ○ | Y |
| PHC 21 | Microfield* | Delivery | Y | N | | | | | N |
| PHC 21 | unknown | Emergency | Y | N | | | | | N |
| PHC 23 | "Axiom" | Delivery | Y | Y | | | | ○ | Y |
| PHC 27 | "VINS"* | Delivery | N[2] | - | | | | | N[2] |
| **TOTAL** | | **N=42** | **28 (67%)** | **17 (40%)** | **9 (21%)** | **2 (5%)** | **1 (2%)** | **5 (12%)** | **5 (12%)** |

Notes: LPM = litres per minute. (1) tested at 5LPM or specified maximum; (2) unable to fully tested due to power outage; (3) missing electrical cable and/or other essential parts; (4) unable to be tested as it was being used for a critically ill patient at the time of survey; (*) had visible Conformité Européenne CE marking indicating compliance with the Declaration of Conformity to ISO 8359.

#### Supplemental table 4: Results from testing of oxygen cylinders in 58 health facilities in Ikorodu local government area, Lagos state, Nigeria

| Facility | Device type | Location | Number | Fit for use[1] |
|---|---|---|---|---|
| **Secondary health facilities** | | | | |
| SHF 1 | Cylinder + manifold | Various[2] | 12 | 12 |
| SHF 2 | Cylinder | Various[3] | ?[3] | ?[3] |
| SHF 3 | Cylinder | Ward (paediatric) | 2 | 2 |
| **Private primary care facilities** | | | | |
| PRV 2 | Cylinder | Delivery | 3 | 0 |
| | Cylinder | Emergency | 1 | 1 |
| PRV 3 | Cylinder | Theatre | 1 | 1 |
| PRV 4 | Cylinder | Triage | 1 | 1 |
| PRV 5 | Cylinder | Theatre | 1 | 1 |
| PRV 6 | Cylinder | Emergency | 5 | 2 |
| PRV 7 | Cylinder | Theatre | 6 | 1 |
| PRV 9 | Cylinder | Ward (adult) | 1 | 0 |
| | Cylinder | Theatre | 2 | 1 |
| PRV 10 | Cylinder | Theatre | 3 | 2 |
| | Cylinder | Clinic | 1 | 1 |
| PRV 11 | Cylinder | Store | 7 | 0 |
| | Cylinder | Emergency | 1 | 1 |
| PRV 12 | Cylinder + splitter | Delivery | 1 | 1 |
| PRV 13 | Cylinder | Emergency | 1 | 1 |
| | Cylinder | Theatre | 1 | 1 |
| PRV 16 | Cylinder | Theatre | 1 | 1 |
| PRV 17 | Cylinder | Theatre | 1 | 1 |
| PRV 18 | Cylinder | Store | 1 | 0 |
| PRV 19 | Cylinder | Emergency | 1 | 1 |
| PRV 20 | Cylinder | Theatre | 1 | 1 |
| PRV 21 | Cylinder | Clinic | 1 | 1 |
| PRV 22 | Cylinder + manifold | Theatre | 1 | 1 |
| PRV 24 | Cylinder | Clinic | 2 | 1 |
| PRV 25 | Cylinder | Store | 2 | 1 |
| PRV 26 | Cylinder | Theatre | 3 | 3 |
| PRV 27 | Cylinder | Delivery | 1 | 1 |
| **Government primary health care facilities** | | | | |
| PHC 1 | Cylinder | Delivery | 1 | 1 |
| | Cylinder | Delivery | 1 | 1 |
| PHC 2 | Cylinder | Store | 1 | 0 |
| PHC 3 | Cylinder | Store | 2 | 1 |
| PHC 11 | Cylinder | Delivery | 2 | 1 |
| PHC 14 | Cylinder | Clinic | 1 | 0 |
| | Cylinder | Delivery | 2 | 2 |
| PHC 15 | Cylinder | Delivery | 1 | 1 |
| | Cylinder | Store | 1 | 1 |
| PHC 16 | Cylinder | Delivery | 1 | 1 |
| | Cylinder | Clinic | 1 | 0 |
| PHC 20 | Cylinder | Delivery | 1 | 1 |
| PHC 23 | Cylinder + splitter | Delivery | 1 | 1 |
| PHC 26 | Cylinder | Delivery | 1 | 1 |
| **TOTAL** | - | - | **82 (100%)** | **53 (65%)** |

Notes: (1) fit for use defined as the number of cylinders with a regulator apparatus or functional outlets from manifold system. (2) Cylinders supplied outlets in 6 wards: Emergency ward, Operating Theatre, Maternity suite, General ward (M and F), and Paediatric ward. (3) Unable to complete cylinder survey at facility – cylinders were available and being used in multiple ward areas and in store, but missing exact location, quantity, and functional assessment.

### Supplemental table 5: Triangulation of pulse oximetry and oxygen access to ward areas in 58 health facilities in Ikorodu local government area, Lagos state, Nigeria

| Facility | Oxygen supply[1] | | | | | | | | Use[2] |
|---|---|---|---|---|---|---|---|---|---|
| | OPD | MAT | ED | GEN | OT | PED | ICU | Store | Oximeter |
| **Secondary health facilities** | | | | | | | | | |
| **SHF 1** | | >2 outlets | >2 outlets | >2 outlets | >2 outlets | >2 outlets 0 concent | >2 outlets | 12-bank manifold | 0 paed* |
| **SHF 2** | cylinder[3] | cylinder[3] | cylinder[3] | cylinder[3] | cylinder[3] | cylinders[3] 0 concentr | cylinder[3] | 25 cylinder | 0 paed |
| **SHF 3** | | | | | | 2 cylinder, 0 concent | | | 1 paed |
| **Private primary care facilties** | | | | | | | | | |
| PRV 1 | | | | | 0 concent* | | | | 0 clinic, 0 OT |
| PRV 2 | | 0 cylinder no gauges | 1 cylinder | | 0 concent | | | | |
| PRV 3 | | | | | 1 cylinder | | | | |
| PRV 4 | | 0 concent | 1 cylinder, 0 concentr | | 0 concent | | | | 1 theatre, 0 clinic* |
| PRV 5 | | | | | 1 cylinder | | | | |
| PRV 6 | | | 2 cylinders, 0 concent | | | | | | 0 clinic*, 0 ED* |
| PRV 7 | | | | | 1 cylinder, 0 concent | | | | 1 OT |
| PRV 8 | | | | 1* concent | | | | | 0 ward* |
| PRV 9 | | | | 0 cylinder no reg | 1 cylinder | | | | 0 clinic* |
| PRV 10 | 1 cylinder | 0 concentr | | 1 concent | 2 cylinder | | | | 1 everwhere |
| PRV 11 | | | 1 cylinder (more in store) | | | | | | |
| PRV 12 | | 1 cylinder + splitter, 0 concent* | | | | | | | |
| PRV 13 | | | 1 cylinder, 1 concent | | 1 cylinder, 0 concent | | | | |
| PRV 14 | | | | 0 concent | | | | | 0 clinic* |
| PRV 15 | | | | | 1 concent | | | | |
| PRV 16 | | | | | 1 cylinder | | | | 1 clinic |
| PRV 17 | | | | | 1 cylinder | | | 0 concent* | |
| PRV 18 | | | | | 0 concent* | | | 0 cylinder no reg | 1 clinic |
| PRV 19 | | | 1 cylinder | | 0 concent* | | | | 0 clinic |
| PRV 20 | 0 concent | | | | 1 cylinder | | | | |
| PRV 21 | 1 cylinder | | | | 0 concent* | | | | |
| PRV 22 | | | | | 1 or more cylinder manifold, 0 concent* | | | | 0 clinic |
| PRV 23 | | | | | 0 concent | | | | |
| PRV 24 | 1 cylinder | | | | 0 concent | | | | 1 gen |
| PRV 25 | | | | | | | | 1 cylinder | 0 matern |
| PRV 26 | | | | | 3 cylinder, 0 concent | | | | |
| PRV 27 | | 1 cylinder | | | 0 concent | | | | 1 OT, 1 clinic |
| **Government primary health care facilities** | | | | | | | | | |

| | | | | | | | | |
|---|---|---|---|---|---|---|---|---|
| PHC 1 | | 2 cylinder, 0 concent* | | | | | | |
| PHC 2 | | | | | | | | 0 cylinder no regulator |
| PHC 3 | | 0 concent* | | | | | | 1 cylinder |
| PHC 4 | | | | | | | | |
| PHC 5 | | | | | | | | |
| PHC 6 | | | | | | | | |
| PHC 7 | | | | | | | | |
| PHC 8 | | | | | | | | |
| PHC 9 | | | | | | | | |
| PHC 10 | | | | | | | | |
| PHC 11 | | 1 cylinder, 0 concent | | | | | | |
| PHC 12 | | | | | | | | |
| PHC 13 | | | | | | | | |
| PHC 14 | | 2 cylinder, 0 concent | | | | | | |
| PHC 15 | | 1 cylinder | | | | | | 1 cylinder |
| PHC 16 | 0 cylinder no reg | 1 cylinder | | | | | | 1 concent |
| PHC 17 | | | | | | | | |
| PHC 18 | | | | | | | | |
| PHC 19 | | | | | | | | |
| PHC 20 | | 1 cylinder | | | | | | |
| PHC 21 | | 0 concent | 0 concent | | | | | |
| PHC 22 | | | | | | | | |
| PHC 23 | | 1 cylinder + splitter, 1 concent | | | | | | |
| PHC 24 | | | | | | | | |
| PHC 25 | | | | | | | | |
| PHC 26 | | 1 cylinder | | | | | | |
| PHC 27 | | 0 concent* | | | | | | |
| PHC 28 | | | | | | | | |

Notes: * Concentrator unable to be tested due to power failure or in use (PRV 8), or pulse oximeter appeared incompatible with Fluke testing device. (1) Adequate oxygen supply for minimum expected use. Numerical values indicate functional oxygen sources located in each ward area ('0' means none of the sources present were functional), while blank cells indicate no oxygen sources in that ward area. Oxygen supply assumptions: secondary health facilities provide inpatient and outpatient services and must be able to simultaneously provide oxygen to 2 patients in each inpatient ward area with backup supply available; private facilities provide outpatient and inpatient services +/- surgical services, and must be able to provide oxygen to one patient in each inpatient ward area with backup supply available; government primary health care facilities provide outpatient and maternity services and must be able to provide oxygen to one patient with backup supply available.; (2) Ability to triage and monitor patients on oxygen with oximetry for minimum expected demand. Numerical values indicate functional oximeters located in each ward area ('0' means none of the oximeters present were functional), while blank cells indicate no oximeter in that ward area. Oxygen use assumptions: pulse oximetry must be available in every ward area that sees acutely unwell patients. (3) Unable to complete cylinder survey at facility – cylinders were available and being used in multiple ward areas and in store, but missing exact location, quantity, and functional assessment. OPD = outpatient department / clinic; MAT = maternity / birth suite; ED = emergency department; OT = operating theatre; PED = children's / neonatal ward; GEN = general / adult wards; ICU = intensive care unit. Red = not available. Orange = available but inadequate to meet needs. Green = adequate for minimum expected service need. * Device not able to be fully tested (e.g. due to power outage).
